# What effects the engagement of splints and orthotics by people after stroke? A qualitative interview study

**DOI:** 10.64898/2026.02.10.26345062

**Authors:** Sarah J Lloyd, Rachel C Stockley

## Abstract

**Background:** Despite recommendations in clinical guidelines, clinical experience indicates that engagement with splints and orthotics varies amongst people after stroke.

**Objectives:** The aim of the study was to understand the factors that influence engagement with splints and orthotics in people after stroke.

**Methods:** People after stroke who had been wearing a splint or orthotic (also known as devices) for at least 2 months under the care of one Community Neurosciences Team in the UK’s National Health Service were included. Semi structured interviews based on the constructs of Bandura’s Social Cognitive Theory (SCT) were used to gather participant’s views, and a framework analysis applying the constructs of SCT was completed using NVIVO software.

**Results:** Four key themes were identified: 1. Self-Regulation; difficulties applying the device and aesthetic acceptability. 2. Self-Efficacy; increased confidence when wearing the device and reduced motivation to wear the device. 3. Outcomes Expectation; reduced falls risk, improved gait, improved balance, maintaining range of movement, and negative effects such as discomfort, pain, itching. 4. Social Support; support needed to apply the device and the burden on family members/carers to apply the device correctly.

**Conclusions:** The findings of this study highlight key factors that influence engagement with orthotics and splints. These include difficulty applying the device after stroke, device aesthetics, comfort, and the importance of continued support from carers. Manufacturers should consider how people after stroke can independently don and doff devices. Education of carers and family members also appears key to support their engagement.

## Introduction

Splints and orthotics (also known as devices) are often used to help manage ongoing limitations in movement after stroke. Splinting is an intervention to prevent and correct contractures and is the process of applying a prolonged stretch through the application of a device [1]. An orthotic is a device which is also applied externally to the body and can be used to improve or support movement and functional impairments such as gait disturbance [2].

Recommendations in clinical guidelines [3,4,5] indicate that splinting should not be offered routinely but after individualised assessment by appropriately skilled staff and specifically following Botox. The use of wrist and hand splints is controversial, some evidence does not support splinting as an intervention for managing spasticity and contractures [6,7]. However, others have demonstrated a positive impact from using wrist and hand splints compared to no splints in range of movement over a 4-month duration [8]. In contrast to splints, orthotics are largely agreed to be beneficial following stroke [9,10], and ankle foot orthoses are recommended in the treatment of foot drop to aid gait re-education [3,4,5].

Despite largely supportive evidence, engagement with splints and orthoses appears variable, with less than half of people after Stroke, Traumatic Brain Injury and Charcot Marie Tooth disease reporting wearing their devices [11,12,13]. However, there is limited evidence of the factors that influence engagement with splinting and orthotics following stroke.

Factors such as discomfort, pain and spasticity have been reported to affect engagement with splints following stroke [11] and the presence and duration of agitation predicted non-compliance to their use after Traumatic Brain Injury [12]. A recent systematic review highlighted that engagement with hand and wrist splints was more likely if there was an immediate benefit, they were comfortable and minimally interfered with lifestyle and daily living [14].

Common barriers to long-term use of lower limb orthotics are reported to include skin irritation, pain, difficulty donning and doffing, appearance and feel, and access to suitably skilled professionals to assess and fit the orthotic [2,15]. In one study, participants stated that benefit and necessity of the orthotics sometimes outweighed the obstacles to wearing them [2]. Patients were more likely to use their orthotics if they had successful outcomes, such as improved gait and balance, activities of daily living and quality of life [2,15].

Understanding the factors that influence engagement with splints and orthotics after stroke is important to guide service delivery, device development and tailor support for users. Consequently, the aim of this study was to evaluate the factors that influence engagement of splints and orthotics in people after stroke by exploring their experiences, and the objective was to identify the barriers and facilitators to engagement of splints and orthotics.

## Material and Methods

### Study Design

The project undertook an interpretative qualitative approach using semi structured interviews. Social Cognitive Theory (SCT) underpinned study design and was chosen to inform the interview schedule as it informs models that promote health and explores health behaviours [16].

Four key constructs were utilised that have been shown to influence self-management in people after stroke [17], namely:

1. Self-efficacy
2. Self-regulation
3. Outcomes expectations
4. Social support

The interview questions were discussed with and reviewed by public advisors to ensure that they were framed appropriately.

### Participants

Participants were purposively sampled community dwelling adults after stroke in the UK who had been provided with a splint or orthotic by a Community Service or other source such as a local Orthotic Department within the National Health Service (NHS) at least two months earlier. Individuals were excluded if they were unable to understand or communicate their needs sufficiently to provide informed consent.

### Procedure

After providing informed consent, participants were allocated a personal identification number (PIN) to enable pseudo anonymisation of all data files and ensure confidentiality and data security.

Participants were then invited to complete a contextual information survey and an interview. Interviews were conducted in the participants home by the project lead (a registered Physiotherapist) and lasted approximately 30 minutes.

The interviewer was a member of the clinical team and to mitigate the risk of bias the interviewer did not recruit participants from their own caseload. In addition, preliminary information on the study was always provide by the participants treating clinician, and the participant was able to have a representative of their choosing present during the consent and interview process.

Ethical approval was achieved from University of Central Lancashire-HEALTH 01090 FR ARC.

### Data Collection

Data collection took place between March and July 2024. A contextual information survey collecting data including age, gender, ethnicity, living and social support arrangements and care package was completed to understand the social determinants of health and health inequalities [18].

Interviews were completed face to face and recorded on a digital encrypted recorder. Interview topics (Table 1) were underpinned by SCT and provided a framework for the interviews whilst allowing flexibility to adapt to individual responses.

**Table 1:** Interview Topics.

|  |  |
| --- | --- |
| 1 | Tell me about the splints/orthotics that you have been given. |
| 2 | <u>Self-Efficacy</u><br>How do you feel about your splints/orthotics?<br>• What makes you feel good?<br>• Do you have any bad feelings towards your splints/orthotics? |
| 3 | <u>Outcomes Expectations</u><br>Tell me about what you feel are the advantages and disadvantages of your splints.<br>• In what way do they benefit you?<br>• In what way do they hinder you? |
| 4 | <u>Self-Regulation</u><br>Can you tell me a bit about your experience of wearing your splints/orthotics?<br>• Was there anything that made it easier?<br>• Was there anything that made it more difficult? |
| 5 | <u>Social Support</u><br>Do you need help with your splints/orthotics?<br>• Who helps you with your splints/orthotics?<br>• How do they help you?<br>• Are you happy to have help with your splints/orthotics? |
| 6 | Is there anything else that you have thought about and would like to tell me? |

### Data Analysis

Recordings were transcribed verbatim, checked for accuracy and pseudo anonymised. Lumivero NVivo 14® software was used to analyse the data. The four constructs of SCT (self-regulation, self-efficacy, outcomes expectation and social support) informed a framework analysis. Codes were developed, categorised, organised and summarised into themes to form a matrix [19]. Data saturation was demonstrated by the completeness and strength of emergent themes and their representativeness across participants [20].

## Results

### Participant Characteristics

Nine community-dwelling adults after stroke participated in the study. Demographic data is displayed in Table 2.

**Table 2:** Demographic Data.

| Participant | Splint/Orthotic | Age Range | Gender | Ethnicity | Housing arrangements | Living arrangements | Care Package |
| --- | --- | --- | --- | --- | --- | --- | --- |
| 1 | Orthotic | 51-55 | Male | White British | Home owner | Lives with family | No |
| 2 | Orthotic | 56-60 | Male | White British | Council renter | Lives alone | 4 calls with 2 carers |
| 3 | Orthotic | 66-70 | Female | White British | Part ownership | Lives with family | No |
| 4 | Splint and Orthotic | 61-65 | Female | White British | Home owner | Lives with partner/spouse | 4 calls with 1 carer |
| 5 | Orthotic | 61-65 | Female | White British | Home owner | Lives with partner/spouse | No |
| 6 | Orthotic | 76-80 | Female | White British | Home owner | Lives alone | 1 carer twice a week |
| 7 | Splint and Orthotic | 51-55 | Male | White British | Private renter | Lives with partner/spouse | 3 calls with 2 carers |
| 8 | Splint and Orthotic | 71-75 | Female | White British | Council renter | Lives alone | No |
| 9 | Orthotic | 61-65 | Female | White British | Home owner | Lives with partner/spouse | No |

### Findings

Key themes for the four constructs of SCT are described below. Table 3 provides an overview of the positive and negative features of each construct to examine the barriers and facilitators.

**Table 3:** Overview barriers and facilitators structured according to Social Cognitive Theory.

| Self-Regulation |  | Self-Efficacy |  | Outcomes Expectations |  | Social Support |  |
| --- | --- | --- | --- | --- | --- | --- | --- |
| Facilitator | Barrier | Facilitator | Barrier | Facilitator | Barrier | Facilitator | Barrier |
| Aesthetically pleasing | Aesthetically unpleasing | Feeling less disabled | Reduced motivation | Comfortable | Uncomfortable | Independence with application | Carer/family burden |
| Easy application | Difficult application | Increased confidence |  | Improved Balance | Dislike | Family/carer support | Carer/family applying device incorrectly |
| Build tolerance to wearing | Application causes fatigue |  |  | Reduced fatigue when walking | Itchy |  | Limitation of care package timings |
| Adjustments made by participant to device to improve engagement | Style and type of footwear |  |  | Improved gait | Perceived non benefit |  | Reduced independence |
| Ideas to improve design |  |  |  | Improved range of movement | Not waterproof |  | Emotional impact of requiring assistance |
|  |  |  |  | Improved rehab outcome | Preferred alternative device |  |  |
|  |  |  |  | Improved alignment of joint/ positioning | Sweaty |  |  |
|  |  |  |  | Improved posture | Too restrictive |  |  |
|  |  |  |  | Reduced falls | Skin Marking |  |  |
|  |  |  |  | Reduces pain |  |  |  |
|  |  |  |  | Increased independence on stairs |  |  |  |
|  |  |  |  | Increased strength |  |  |  |
|  |  |  |  | Provides stretch |  |  |  |
|  |  |  |  | Provides support |  |  |  |

#### 1. Self-Regulation

Two clear themes emerged: aesthetics and the application of the device.

Some participants described positively how the device was discreet and therefore not detectable by the general public:

> *“you can’t see it, you don’t, nobody would look at you and know you had got it on. So, you just feel comfortable with it on*.*”* (Participant 5)

However, one participant described their device as aesthetically displeasing indicating that wearing it in public would make them less likely to engage with their device:

> *“I wouldn’t want to wear it going out in public really*.*”* (Participant 7)

Application of the device was frequently discussed as participants described how weakness in their upper limb made it challenging to apply splints and orthotics independently:

> *“I can’t get it on properly. With two hands it wouldn’t be a problem. But with the one hand, no, that is the only thing I have got against it, that I can’t actually get it on properly myself*.*”* (Participant 6)

In addition, participants reported that the effort and fatigue involved in applying the device resulted in them being less likely to wear it.

> *“I have tried to think my way through it. And I have achieved it but to do it just to go from here to the loo, it is easier just to sort of not use it you know*.*”* (Participant 7).

However, some participants reported that they found it easy to apply their device and that with practise this improved with time.

> *“Yes, I find it easy. Not at first but yes you just get used to it*.*”* (Participant 5).

Participants acknowledged that although the device could be challenging to apply it was worth the effort:

> *“it is not a quick thing that I can rush round and do. It does take time, but it is worth it*.*”* (Participant 9)

Some participants described how they developed adjustments to make the device more tolerable to wear or improve ease of application. Modifications included:

1. Using a stool to reduce bending down
2. Using talcum powder to reduce irritation
3. Permanently fixing parts of orthotic to trainer
4. Using a sock as a barrier to reduce irritation

Other participants expressed ideas to improve the design of devices to make them easier for people after stroke to use, particularly in relation to single handed devices:

> *“I don’t think it would be too much to come up with a design to actually get it to be a one-handed thing you know*.*”* (Participant 7)

#### 2. Self-Efficacy

Participants consistently reported that using their devices made them feel more confident, this predominantly related to walking and reduced falls:

> *“It gives me the confidence to know that I have got something supporting that foot, and hopefully with that on, I won’t catch my foot and drag my foot and fall*.*”* (Participant 6)

One participant described feeling “less disabled” when using their device. This was in relation to the position of their arm when the device was in place:

> *“When like I am sat here now and I can feel that [bent], I feel like, I feel a bit disabled and I don’t like feeling disabled*…*whereas when that’s on its straight all the time*.*”* (Participant 8)

In contrast, one participant felt strongly that they did not want to use their device and was not motivated to wear it:

> *“I didn’t want it on and that was it. I couldn’t be bothered with it*.*”* (Participant 2)

#### 3. Outcomes Expectations

Many participants described the positive effects of their devices:

> *“I don’t think I would be as far as I am now without it*.*”* (Participant 8)

In addition to feeling more confident as highlighted within self-efficacy participants noted improvements in their functional recovery such as improved gait, increased balance and reduced falls:

> *“It is easier for me to walk. My leg is not dragging, and my foot is not dragging. Like for instance, during the night, if I go to the bathroom without it on, my foot tends to drag and my toes curl under. Whereas when I have this on, it is fine, it feels okay*.*”* (Participant 9)

Other functional activities such as performing the stairs were improved when wearing the devices, enabling participants to be more independent around the home:

> *“I will always have it on when I go up the stairs, I can ascend and descend the stairs independently with it on now*.*”* (Participant 7)

Participants felt that the devices provided extra support and resulted in improvements with posture and alignment. Some participants described how the devices had a strengthening effect, whilst others reported an improvement in range of movement and stretch, however some noted that the devices could limit function by being too restrictive.

> *“when it is on I can’t particularly use my hand*.*”* (Participant 3)

Participants reported that the devices were largely comfortable:

> *“It straightens my foot up a little bit so that when I am putting the foot down, it is much more comfortable and it has just got a little padding underneath so that, that helps with my toe, which curls under. So, it just feels nice and secure, nice and comfortable and I can walk round in it and not even know I have got it on*.*”* (Participant 5)

However, they did describe unfavourable side effects including discomfort, itching, sweating and skin marking which resulted in reduced tolerance of the devices:

> *“It was like skin irritation because it gets a little bit sweaty*.*”* (Participant 7)
>
> *“I will take it off and have a couple of hours. Because my foot does go red, my foot does go red when I have been wearing it*.*”* (Participant 10)

#### 4. Social Support

Independence with application of the splint or orthotic was a key facilitator to participants engaging with their device.

Participants reported negative connotations of requiring assistance to don and doff the device, discussing a reluctance to ask for help to apply the device, finding it embarrassing, and the lack of independence frustrating:

> *“I wish I could do it myself. Because you feel, after a stroke I think you feel you have lost all your independence, you can’t do anything yourself so I really would prefer to do things myself but it’s just impossible, I just wouldn’t be able to do it”*. (Participant 5)

Similarly, participants reported positive effects of being able to apply the device without help:

> *“I can do that myself, which is much easier, I don’t have to ask anyone”*. (Participant 5)

Although some participants were happy to receive support from carers or family, others described that they were reliant on family members or carers to apply the devices which increased carer burden.

In addition, one participant described a variation between carers in the consistency of applying the device which influenced its effectiveness:

> *“There is inconsistency with my hand splint. Some people could put it on and other people can’t”. “Some carers can do it quite easily and others again they struggle*.*”* (Participant 3)

Finally, one participant described the barriers of care package timings and how this effected engagement as the participant could not tolerate having the device on for the length of time between care calls:

> *“They [the carers] did put it on sometimes, and it was only supposed to be on until 12 o’clock and sometimes the carers didn’t come back till b***** 1 o’clock something like that, or sometimes later. 3 o’clock I had it on till once. I took it off myself”*. (Participant 2)

## Discussion

This qualitative study provides important insight into the factors that could affect engagement of splints and orthotics in people after stroke. Key findings highlighted that people after stroke valued devices that were discreet, aesthetically pleasing, and easy to don and doff independently. Participants consistently reported that devices made them feel more confident and described functional benefits such as improvements with gait, balance and falls alongside postural improvements with support and alignment. Comfort of the appliance was a key theme, with unfavourable side effects such as pain and itching negatively affecting use, whilst comfortable devices were seemingly more tolerable. Participants noted that having support to don and doff the device was often needed due to weakness from the stroke, which was a source of frustration. The presence of a carer who could help with application of the device could facilitate use but added to carer burden and required skills were variable.

Barriers to use of splints and orthotics including ease of application, aesthetic appearance of devices, comfort, and reliance on carers/family members have been highlighted by others [2,11,14,15] supporting this finding. However, in this study participants also described how the capability of family members and carers to don and doff devices, and suitable care package timings affected engagement with devices, generating additional understanding of wider factors influencing engagement of these devices in the community.

In keeping with Bandura [16], participants in this study indicated that positive experiences (outcomes expectations) and self-efficacy influenced their willingness to use devices. Those participants who perceived that the devices made them more confident, improved their gait, balance and falls risk, or the position of a joint, had a positive attitude to using their splints and orthotics which has also been reported in other studies [2,14,15]. In contrast, participants who perceived little benefit of the device or who had negative connotations towards the device were seemingly less likely to use them, which has been conveyed by others [2,11,13,14,15].

### Implications for practice

This study has highlighted several key areas which could influence engagement with device use after stroke. A clear finding was that devices that can be applied using one hand and are discreet and aesthetically pleasing to people after stroke appear more likely to be used consistently. Producers of splints and orthotics could collaborate with those who have lived experience of stroke during the design and manufacturing process of devices to ensure usability and longevity. In other areas of stroke rehabilitation, co-design has been used with success, including gait assessment tools and digital health platforms [21,22]. Manufacturers could also consider how devices can be applied in different styles of footwear offering flexibility to people after stroke when using their devices and increasing engagement.

The importance of understanding the potential benefits of devices was highlighted in this study suggesting that education and training for people after stroke and their carers could be beneficial. Areas such as considering the advantages and disadvantages of devices, wearing times and problem-solving methods of donning and doffing devices could be included in a training package. These packages should be provided in variable formats (including written, photographic and/or video presentations) and co-developed with people after stroke and carers to have maximum impact. The positive benefits of carer training and education have been well reported [23,24] and include reduced care costs, carer burden and patient and carer anxiety and depression, with improved patient and carer quality of life and satisfaction of rehabilitation programs. Regular reviews of devices to address unfavourable side effects and difficulties using their splints and orthotics could also be incorporated in a follow up pathway which may help maintain people after strokes motivation and engagement. Consideration should be given to providing recommendations that work in conjunction with care package timings and/or liaising with care agencies to alter care package timings to suit service users as described by participants in the current study.

### Limitations of the study

The relatively small sample and limited diversity of participants interviewed for this study is a clear limitation to its generalisability to the wider stroke population. However, data saturation was achieved, suggesting consistency in findings of this study. It should also be noted that splints and orthotics were discussed collectively and there is a risk that participants may have based their answers on one device if they had been provided with both.

### Future Research

Future research studies could endeavour to recruit a broad spectrum of ethnicities to understand whether there are any further barriers, facilitators or health inequalities affecting engagement with devices within this demographic. Further work could also investigate whether there are any differences to engagement with devices during acute inpatient and community phases of rehabilitation. A national multi-centre study could be developed to identify the extent of engagement of splints and orthotics in people after stroke across the country, this would give a broader awareness of any issues.

## Conclusion

The study successfully achieved its aim of gaining a deeper understanding into the factors that influence engagement with splints and orthotics in people after stroke. Key findings highlighted that people after stroke valued devices that were discreet, comfortable, aesthetically pleasing, and easy to don and doff independently. Requiring support to apply a device was reported as a barrier to engagement.

Manufacturers of splints and orthotics would benefit from working with people after stroke who have lived experience of splints and orthotics when designing devices. Education of people after stroke, family and carers is key for support, application and engagement of devices, and therapists should take an individualised approach to each service user investigating their personal barriers and facilitators to engagement of devices.

## Data Availability

All data produced in the present study are available upon reasonable request to the authors

## Data availability

The data that support the findings of this study are available from the corresponding author upon reasonable request.

## Acknowledgements

The authors would like to thank the Physiotherapists and Occupational Therapists within the Community Neurosciences Team who assisted with participant recruitment. They would also like to thank the participants who gave their time to contribute to the study.

